# Prevalence of Iron, Vitamin B12 deficiency and inflammatory anaemia in treatment naive individual consecutive cancer patients: A cross sectional study

**DOI:** 10.1101/2020.07.29.20164426

**Authors:** A Pandey, S Singh, R Aryan, K Murari

**Affiliations:** Department of Medical Oncology, State Cancer Institute, Indira Gandhi Institute of Medical Sciences, Patna, Bihar, India

**Keywords:** Iron deficiency, B12 deficiency, Inflammatory Anaemia, Cancer Anaemia

## Abstract

**Background:** In treatment naive Indian cancer patients, prevalence of iron, B12 deficiency and inflammatory anaemia in poorly known.

**Aims and Objectives:** To evaluate prevalence of anaemia and iron, B12 deficiency along with inflammation in treatment naive individual consecutive cancer patients.

**Material and Methods:** All patients registered from 1st July 2019 till 31st December 2019 in Medical Oncology Outpatient Department were offered to undergo Iron profile, Serum B12 levels and Serum ferritin along with routine haematological investigations. Anaemia was defined as Haemoglobin < 11gm/dL. Transferrin saturation <20%, Serum Ferritin >300 microgram/litre and Vitamin B12 level <200 picogram/millilitre were ‘cut-offs’ used to define iron deficiency, inflammation and Vitamin B12 deficiency respectively. Data was analysed using descriptive statistics, frequency distribution, crosstabs and Bar Diagram in SPSS version 17.0. Pearsons Chi square test and Odds ratio was used to measure the strength of association with variables.

**Results:** 311/441 (70.5%) were found eligible. Median age was 52 ± 15.9 (Range 18-84) years with 144/331(46%) females. The prevalence of anaemia was 61% ± 2.7 (95% CI 55-66%). Mean Haemoglobin was 9.86 ± 2.08 (range 3-16) gram/decilitre. 21/311(7%) had severe anaemia (Haemoglobin < 6.9 gm/dl). 135/311 (71%), 61/189 (32%) and 89/189 (47%) anaemic patients had iron deficiency, inflammation and B12 deficiency respectively. More than 70% of Gastrointestinal (50/69), Gynaecological(17/24) and Lung Cancer(18/22) patients had underling Iron deficiency.

**Conclusion:** Two-third of cancer patients are iron deficient. B12 deficiency and inflammation are present in half and one-third patients respectively.

## Introduction

Higher prevalence of anaemia in cancer is associated with inferior outcomes, including poor survival and inferior quality of life.^1–3^ Blood loss, malnutrition, haemolysis, chronic inflammation, bone marrow suppression or decreased erythropoeisis, all can individually or in unison cause drop in haemoglobin among cancer patients.^3,4^ Treatment with chemotherapy or radiotherapy can further compromise red blood cell mass and advanced stage tumours have often higher prevalence of anaemia.^4,5^ Further, distribution of anaemia among cancer patients may vary considerably across different countries and hence need to be studied individually, specific to local population cohort.^6–8^

Most of the studies published have primarily focussed on iron deficiency component of cancer anaemia and subsequent remedial therapeutic options, including iron supplementation and blood transfusions.^2,6–8^ A more comprehensive effort with other possible co-variables, such as vitamin deficiencies and chronic inflammation apart from iron deficiency is merited to discover other salient but inconspicuous causes of pre-treatment cancer anaemia, preferably prospectively. We report an Indian prospective cross sectional study among treatment naive, newly diagnosed adult individual consecutive patients across several subtype of malignancies and report underlying iron, vitamin B12 deficiency and chronic inflammation, irrespective of haemoglobin status.

## Material and Methods

### A. General Study details

This is a prospective cross-sectional study of individual consecutive treatment naive adult cancer patients registered in Department of Medical Oncology at Regional Cancer Centre, Indira Gandhi Institute of Medical Sciences, Patna from 1st July, 2019 till 31st December 2019. It is a non interventional, observational study with aim to illicit the prevalence of anaemia, Iron-B12 deficiency and chronic inflammation before receiving any cancer directed therapy. Written informed consent was obtained from each participant before enrolment. Apart from routine pre-treatment work up investigations such as Complete Hemogram, serology, coagulation profile, renal and hepatic function tests, we also obtained peripheral venous blood samples for Serum Iron, Serum Ferritin, Total Iron Binding Capacity, transferrin saturation and serum B12 levels. The study was conducted in accordance with Declaration of Helsinki and the Indian Council of Medical Research (ICMR) guidelines for ethical conduct.^9^ No funding was received for the conduct of the study.

### B. Eligibility Criteria

All adults of age more than 18 years, irrespective of type of malignancy were eligible for this study. Patients enrolled should have cytology, biopsy, serological, tumour markers or flow cytometry based confirmation of malignancy and its type either at the time of enrolment or soon after (within 2 weeks). Any patient who has received prior anti-cancer therapy including chemotherapy, hormonal, radiotherapy or major surgery, except diagnostic biopsy/ cytology were excluded. Patients found pregnant, with chronic renal failure failure, decompensated liver cirrhosis, inflammatory bowel diseases, history of major gastric or bowel resection, active life threatening infection, chronic active infection e.g Tuberculosis, Leishmaniasis, active rheumatological or flared up connective tissue disorders such as Rheumatoid arthritis, spondyloarthropathies, Systemic Lupus Erythromatosis, scleroderma or sarcoidosis were excluded.

Patients with documented psychiatric condition interfering with diet such as anorexia nervosa or bulimia were excluded. Any patient with history of prior prescription of oral / parenteral iron or B12 supplementation in past three months were not considered. Any history of blood transfusion in past one month were excluded. Patients with history of chronic gastrointestinal bleed, including any tumour related events such as bleeding per vaginum or hemoptysis, hematemesis or heamaturia were however eligible to participate.

### c. Sample collection and processing

‘Three milliliters blood sample was drawn from an accessible peripheral vein in aplastic tube with or without gel barrier [red or yellow-top serum separator vacutainer] irrespective of fasting or special dietary requirements. Serum was removed from the red blood cell clot and stored at 2°C–8°C refrigeration, if the sample could be processed on the same day or was frozen and stored at −20°C if processing was delayed. Care to be taken to exclude any sample with gross hemolysis, fibrin, or particulate matter. Samples once thawed were mixed thoroughly by low-speed vortexing or by inverting ten times. Specimens were centrifuged for 3000 rpm for 10 min in centrifugation tubes to maintain consistency in results. Only clear samples withoutbubbles were transferred after centrifugation for further processing. For sample processing, Glass tubes were not tested. The average turnaround time for reports was 24–48 hours.

a. For Serum Iron estimation: The MULTIGENT iron assay was used which is intended for the direct colorimetric determination of iron without deproteinization in human serum on the ARCHITECT cSystems. The iron Assay is standardized against National Institute of Standards and Technology Standard Reference Material 3126
b. For Serum Ferritin Levels: The ARCHITECT Ferritin assay was used which is a chemiluminescent microparticle immunoassay [CMIA] for the quantitative determination of ferritin in human serum onARCHITECT iSystems. The Ferritin Assay is standardized against the World Health Organization (W.H.O.) Ferritin First International Standard 80/602
c. For Transferrin: The Transferrin assay was used which is an immunoturbidimetric procedure for determination of transferrin in human serum on the ARCHITECT cSystems. The Transferrin Assay is standardized against reference method calibrated against ERM-DA470.80/602.
d. For serum B12 levels: The ARCHITECT B12 assay was used which is a Chemiluminescent Microparticle Intrinsic Factor assay for the quantitative determination of vitamin B12 in human serum on the ARCHITECT iSystem. Abbott manufactures B12 internal standards gravimetrically using Cyanocobalamin (USP Reference Standard). The B12 calibrators are manufactured and tested against these internal standards.

### D. Data collection, sample size and Definition

Demographic data including age, sex, type of malignancy, stage were recorded. Anaemia was classified as per definition of Indian Council of Medical Research (ICMR) into Mild Anaemia (Haemoglobin 10.9 −10 gram/decilitre), Moderate Anaemia (Haemoglobin 9.9-7 gram/decilitre), Severe Anaemia (Haemoglobin 6.9-4 gram/decilitre), Very Severe (Haemoglobin less than 4 gram/decilitre). Anaemia was also classified as per Mean Corpuscular Volume into Microcytic (< 79.9 femtolitres), Normocytic (80-100 femtolitres) and Macrocytic (>100.1 femtolitres) anaemia. The formal sample size calculation was not done and all the patients in the defined time period from 1st July 2019 till 31st December 2019 were assessed for eligibility into the study. Patients for whom iron profile and B12 level estimation was ordered but the reports were not traceable due to technical failure or not reported (n=18) were classified as data Missing completely at random (MCAR). This missing data was subject to Listwise deletion for complete case analysis making the final sample size of 311. As a result of this, the estimated parameters including prevalence of anaemia are not biased by the absence of the such missed data.

To maintain uniformity in methods of assessment we defined Iron deficiency as those patients having transferrin saturation less than 20%. Patients were classified to have inflammatory state if their serum ferritin levels were more than 300 microgram/litre. Vitamin B12 was defined as low when serum B12 levels were less than 200 picogram/millilitre. All patients were categorised as iron deficient, B12 deficient and in inflammatory state irrespective of having anaemia or not, using above definitions.

### E. Patient Management Protocol

Our threshold for Packed cell transfusion was less than 7.0 gram per decilitre of haemoglobin, unless impending cardiac, renal morbidity, major active bleeding, clinical decision or sepsis merited to reduce transfusion threshold. At the time of recording haematological reports (within two weeks of registration), including complete haemogram, iron profile and serum B12 levels, patients who were anaemia and had Iron or B12 deficiency were given option to receive supplementation. Patients were prescribed either intravenous injectable ferric carboxymaltose (1000 mg flat dose) or ferrous sulfate capsules (100 mg) three times a day for a month. None of the patients were prescribed Erythropoeitin Stimulating Agents (ESA). Similarly for B12 deficient anaemia, 1000 microgram of cyanocobalamine intramuscular injection once daily for seven days, then weekly for one month, then monthly for 6 months were prescribed. However, patients were not followed to check compliance or change in haemoglobin, as this was beyond the purview of study. We did not define or measured any clinical, haematological or serological endpoint for later follow up, irrespective of deficient store replenishment status, as this was a single point cross sectional study of prevalence only.

### F. Statistics

All data were recorded in real time in the Outpatient Department of Medical Oncology by a trained data entry operator under the supervision of the physician. Descriptive statistics, including tables, Venn diagram, charts and frequency distribution, were derived from SPSS software version 17.0 (IBM Corp. Released 2017. IBM SPSS Statistics for Windows, version 17.0. Armonk, NY, USA). Pearsons Chi Square test was used to analyse association of anaemia with several relevant categorical variables. Odds ratio was calculated to measure the strength of association, if any using above software.

## Results

### A. Patient characteristics

Out of 441patients screened and offered baseline Iron profile and serum B12 estimation, only 311 were finally enrolled in the study. [see FLOW diagram, Fig1]. Median age at diagnosis was 52 ± 15.9 (range 18-84). 167 (54%) were males. 79% had advanced stage III/IV cancers and 19% had haematological malignancies. Mean Haemoglobin of entire population was 9.86 ± 2.08 (range 3-16). See Table 1.

**Table 1.** Demographic variables of patients enrolled in the study.

| Sr. No | Variables | Values |
| --- | --- | --- |
| 1. | **Age in years** (median) | 52 ± 15.9 ( range 18-84) |
| 2. | **Sex** |  |
|  | Male | 167 (54%) |
|  | Female | 144 (46%) |
| 3 | **Mean Haemoglobin** (gram/ decilitre) | 9.86 ± 2.08 (3-16) |
| 4 | **Haemoglobin Indices** |  |
| | Mean Corpuscular Volume (MCV in femtolitre) | 87.6 ± 9.96 (61-126) |
| | Mean Corpuscular Haemoglobin Concentration (MCHC in gram/decilitre) | 28.34 ± 7.55 (16.5 -39) |
| | Mean Corpuscular Haemoglobin ( MCH in picogram/cell) | 31 ± 4.64 (20-39) |
| 5 | **Iron Indices** |  |
| | Mean Serum Iron (microgram/decilitre) | 50.53 ± 29.8 (5.5-208) |
| | Mean Total Iron Binding Capacity ( microgram/decilitre) | 285.30 ± 99.2 (59-547) |
| | Mean Serum Ferritin levels (microgram/litre) | 296.41 ± 57.8 (6.9-2131) |
| | Mean Transferrin saturation (%) | 19.04 ± 11.34 (2.5-63.1) |
| 6 | **Mean Vitamin B12 levels** (picogram/millilitre) | 299.93 ± 99.14 (23-907) |
| 7 | **Primary Site/ Type of Malignancies** |  |
|  | Head Neck | 12 (3.9%) |
|  | Genitourinary | 20 (6.4%) |
|  | Central Nervous System | 2 (0.6%) |
|  | Breast | 59 (19%) |
|  | Lung | 22 (7.1%) |
|  | Gastrointestinal | 69 (22.2%) |
|  | Gall Bladder | 35 (11.3%) |
|  | Gynaecological | 24 (7.7%) |
|  | Haematolymphoid | 59 (19%) |
|  | Sarcomas | 7 (2.3%) |
|  | Miscellaneous | 2 (0.6%) |
| 8 | **Stage of Cancer** |  |
|  | I | 4 (1.28%) |
|  | II | 26 (8.36%) |
|  | III | 105 (33.7%) |
|  | IV | 140 (45%) |
|  | Not applicable | 36 (11.5%) |

### B. Anaemia profile

The prevalence of anaemia in treatment naive cancer patients was 61% ± 2.7 (95% CI 55-66%). [See Table 2]. 135/311 (71%), 61/189 (32%) and 89/189 (47%) anaemic patients had iron deficiency, inflammation and B12 deficiency respectively. See [Table 3]. The prevalence of anaemia across several major sites/types of cancers was 71% (42/59) in haematolymphoid, 58% (40/69) Gastrointestinal malignancies,59%(35/59) Breast cancer,68% (24/35) Gall bladder cancer, 66% (16/24) Gynaecological cancer and 50% (11/22) in Lung cancers. More than 70% of Gastrointestinal (50/69), Gynaecological(17/24) and Lung Cancer(18/22) patients had underling Iron deficiency. [See Fig 2]. There was considerable overlap with patients having concurrent iron deficiency, B12 insufficiency and chronic inflammation. 16/311 (5%) of patients had all above three combined. See Venn diagram [see Fig 3]

**Table 2.**
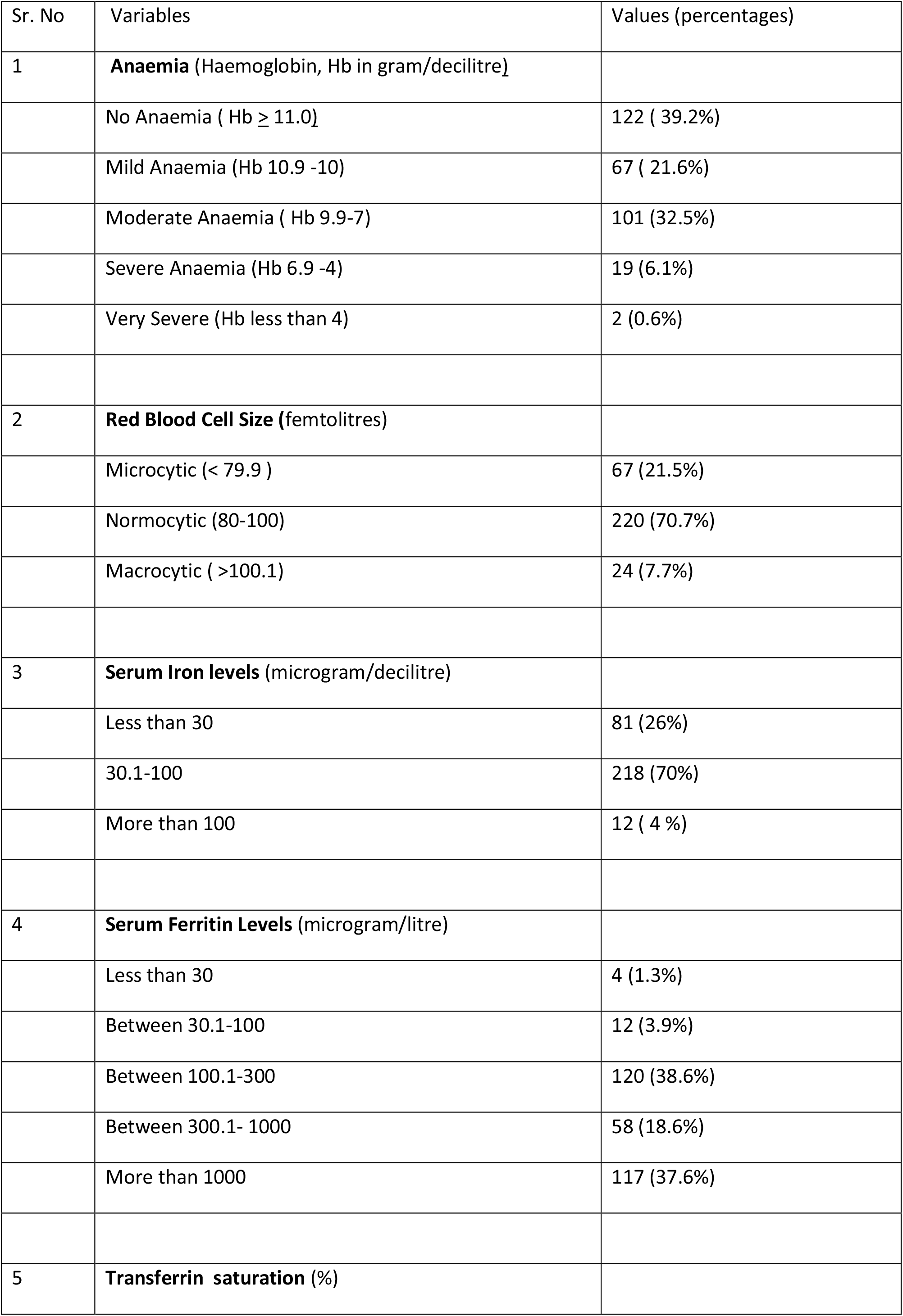
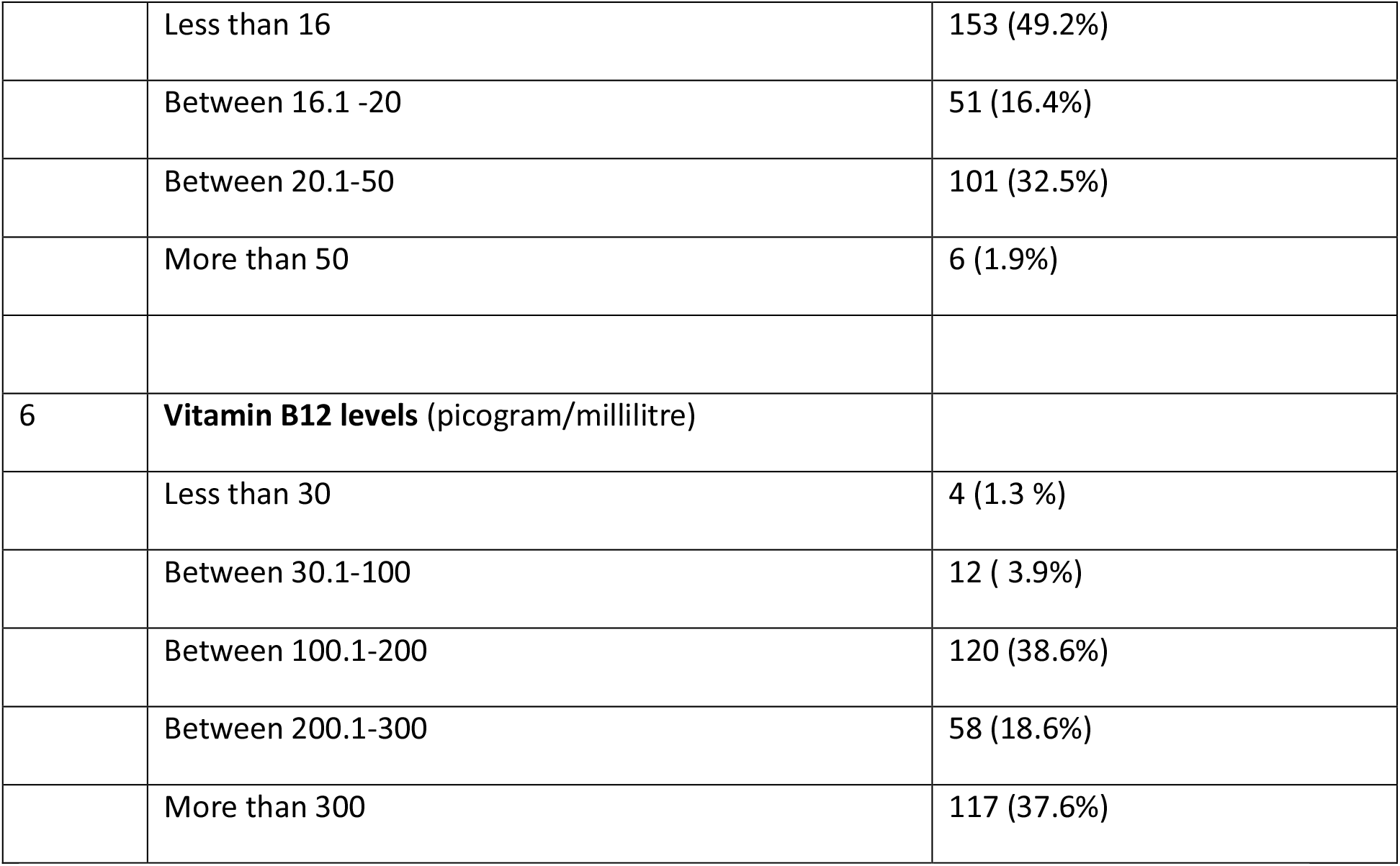
Anaemia and corresponding laboratory variables including iron profile and B12 values.

**Table 3.** Anaemia status with respect to Iron deficiency, chronic inflammation and Vitamin B12 serum levels.

| Sr. No | Anaemia | Iron deficiency<br>(Transferrin<br>saturation less<br>than 20 %) | Inflammatory<br>(Serum Ferritin<br>more than 300<br>microgram/litre) | Low Vitamin B12<br>levels ( Serum B12<br>less than 200<br>(picogram/millilitre) |
| --- | --- | --- | --- | --- |
| 1 | No anaemia (n=122) | 69 (56%) | 26 (21%) | 47 (38%) |
| 2 | Mild Anaemia (n=67) | 43 (64%) | 19 (28%) | 25 (37%) |
| 3 | Moderate Anaemia (n=101) | 76 (75%) | 35 (35%) | 49 (48%) |
| 4 | Severe Anaemia (n=19) | 14 (74%) | 6 (43%) | 13 (68%) |
| 5 | Very Severe (n=2) | 2 (100%) | 1 (50%) | 2 (100%) |
| 6 | **Total \* (n=189)** | **135 (71%)** | **61 (32%)** | **89 (47%)** |

**Figure 1.**
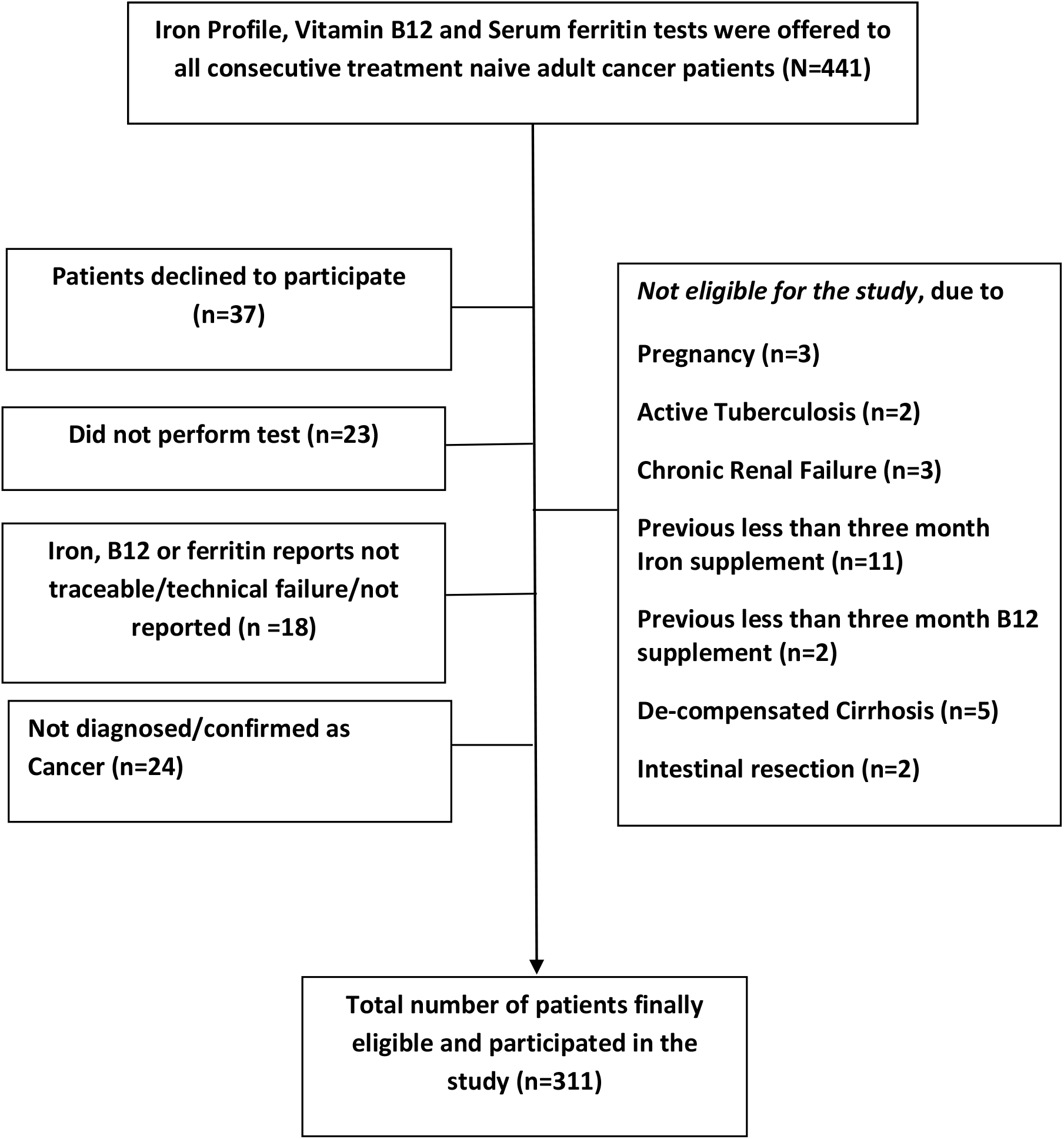
FLOW diagram showing the flowchart of participants accrual and eligibility for the study.

**Figure 2.**
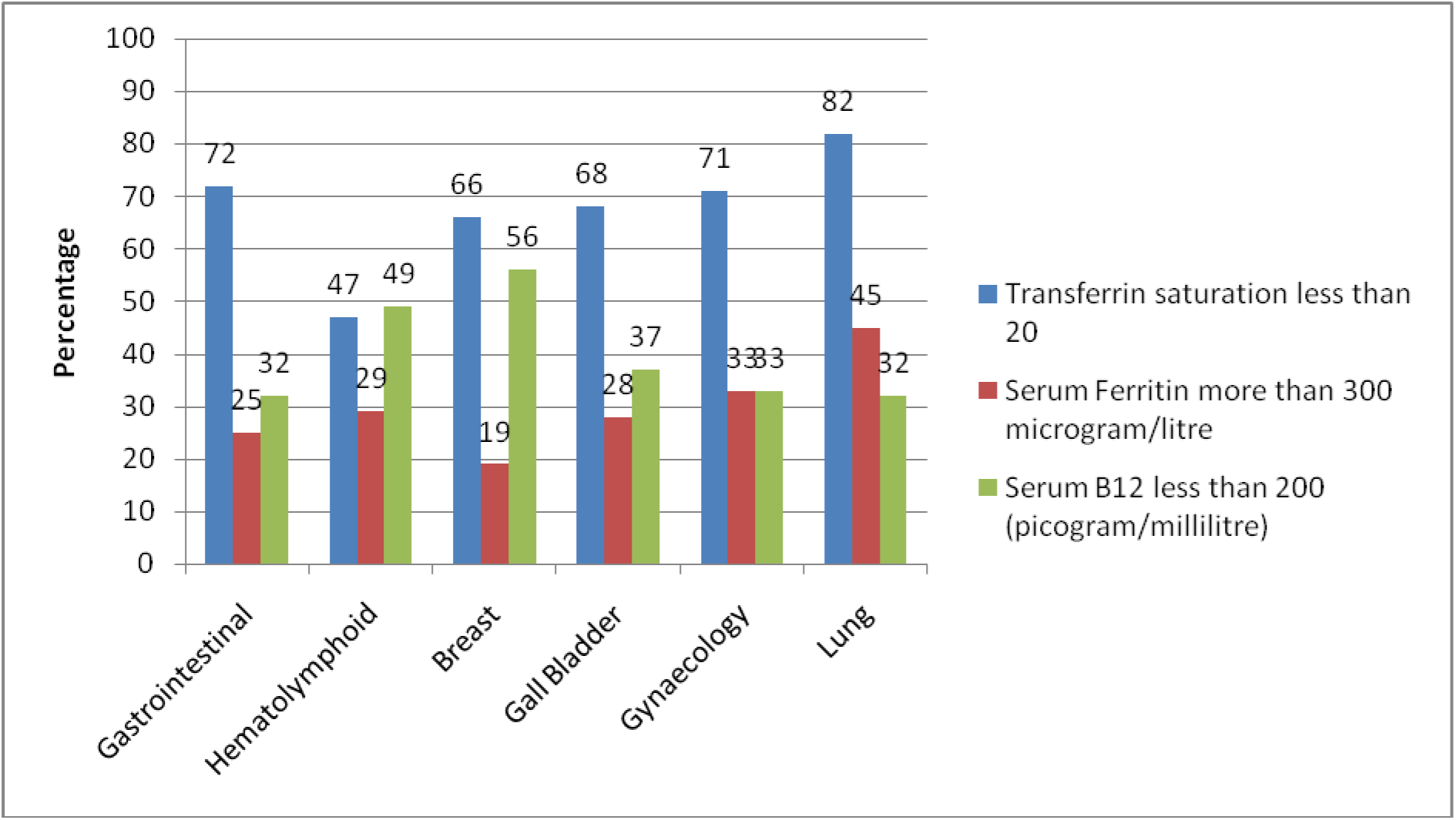
Percentage of patients with Iron deficiency, inflammation and B12 deficiency in several cancer types/sites.

**Figure 3.**
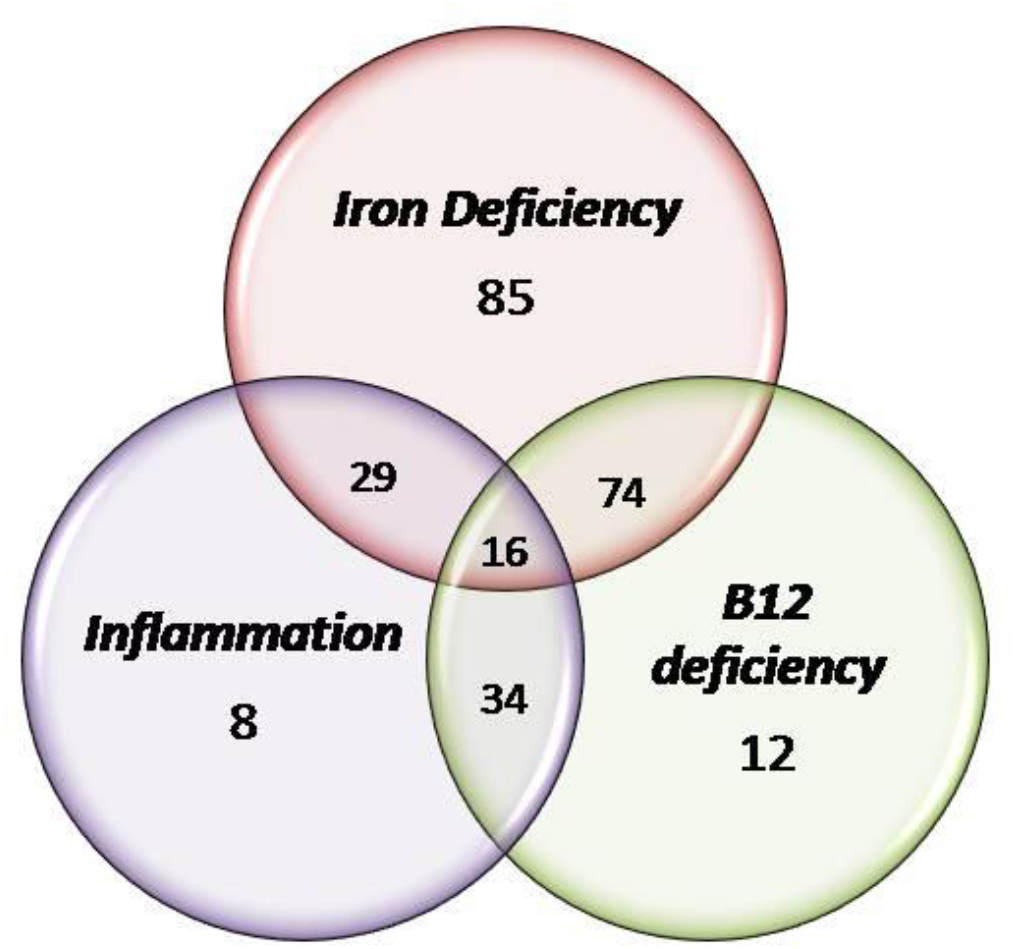
Venn Diagram of Serum Iron, B12 deficiency and Inflammation showing overlap among the three entities.

### C. Association of variables with anaemia

Iron deficiency was strongly associated with anaemia with Pearson Chi square value of 8.05 (p =0.005) and Odds ratio of 1.98 (95% CI-1.23-3.195). Similarly, chronic inflammation was significantly associated with anaemia with above value as 4.13 (p=0.042) and Odds ratio of 1.72 (95% CI-1.02-2.90). Females had higher probability of anaemia with Pearson Chi square value of 4.90 (p=0.027) and Odds ratio of 1.67 (95% CI-1.06-2.65) compared to males. Underlying B12 deficiency was not associated with anaemia in terms of statistical significance with Pearson value of 2.48 (p = 0.115) and Odds ratio of 1.45 (95% CI 0.91-2.30). Similarly, neither higher age (more than 60 years or less) nor type of malignancy (haematological or solid cancers) had shown significant association with respect to anaemia status.

### D. Supplementation for anaemia

Out of 21 patients with haemoglobin less than 7 gram per decilitre, blood transfusion was administered in 19 (90%) within two weeks of getting registered, while one patient declined and another defaulted. 122/135 (90%) were prescribed iron supplementation, while nine declined, three defaulted and one died by second week of registration. Among patients who received iron supplementation, 84/122 (69%) received intravenous iron along with intravenous chemotherapy, while 38/122 (31%) preferred oral iron capsules. Only 58/89 (65%) with B12 deficient anaemia received intramuscular cyanacobalmine for at least seven days of prescription, while 19/89 (21%) received less than seven days and another 12 declined for it.

## Discussion

We report the results of our prospective cross sectional study to evaluate the prevalence of anaemia in adult treatment naive individual consecutive Indian cancer patients. The prevalence of anaemia was 61% ± 2.7 (95% CI 55-66%) with mean haemoglobin of entire population of 9.86 ± 2.08 (range 3-16). 71%, 32% and 47% anaemic patients had underlying iron deficiency, inflammation and B12 deficiency respectively. More than 70% of Gastrointestinal, Gynaecological and Lung Cancer patients had underling Iron deficiency. As per the best of knowledge of authors, this is the first prospective cross sectional study in individual consecutive adult patients across several types of malignancies from India to report the prevalence of underlying pre-treatment anaemic status.

In one of the largest prospective observational study conducted in twenty four European countries with 15367 cancer patients spread across 248 centres, the prevalence of anaemia at enrolment before cancer directed therapy was 31.7%.^10^ They defined anaemia as haemoglobin less than 12.0 gram per decilitre as per American Society of Clinical Oncology and Common Toxicity Criteria, National Cancer Institute(NCI)^11,12^. They showed presence of anaemia in more than 70% of patients with lung cancers or Gynaecological malignancies either at baseline or at some time point within six months of enrolment, including treatment period.^10^ In another observational study reported from Italy and Austria, with Haemoglobin cut off of 10.0 gram per decilitre, the prevalence of anaemia was 32%.^7^ Majority of patients (80%) had solid malignancies and 13 % had used Erythropoeitin Stimulating Agents (ESA) for resolution of symptoms. Both above studies, also mention use of Iron (oral or intravenous) supplementation apart from blood transfusion and ESA for patients with anaemia. However, none of the above two large studies report the baseline iron stores, serum ferritin levels as marker of inflammation or serum B12 levels to establish the exact etiology and proportional role of above three elements in pathophysiology of anaemia in treatment naive cancer patients. There is paucity of similar comprehensive observational study evaluating prevalence of anaemia in treatment naive cancer patients from Indian subcontinent.

Anaemia prevalence varies widely across different continents and countries. World Health Organisation (WHO) estimates the prevalence of anaemia among general healthy population as 23% in Europe, while 45-47% among Africans and South East Asians.^13^ In India, the prevalence of anaemia also differs among different high risk groups such as Pre-school children (74%), School girls (50%) and pregnant women (52%).^13,14^ Hence, it is pertinent to have country or state specific prospective study to illicit anaemia prevalence, especially among high risk group such as newly diagnosed cancer patients. Few attempts have been made across various cancers in India and reported prospectively. In a study done among 96 newly diagnosed and treatment naive lung cancer patients reported from single centre in South India, the anaemia prevalence was 61.4%.^15^ In another large prospective study done exclusively in newly diagnosed lymphoid malignancies with 316 patients recruited over eighteen months, the prevalence of anaemia was 42.4 %.^16^ Nutritional deficiency as the cause was seen in less than half of anaemic cancer patients.

In the Regional Cancer Centre(RCC) of our institute, medical oncology department is a single faculty and single point of contact for all solid and haematological malignancies registered for receiving chemotherapy.^17^ This provide a unique vantage point with ‘bird’s eye view’ across several subtypes of cancer treated under one roof, unlike many apex cancer centres where site and sub-site super-specialization conforms to restricted and diverse disease management patient pool. Our prospective observational cross sectional study includes 311 individual consecutive adult cancer patients of all major sites/ types of cancer. We report the prevalence of anaemia as 61%. This is in accordance with our previous published prospective study in treatment naive cancer patients, where prevalence of Vitamin D deficiency was 67%.^18,19^ Both these highlight poor nutrition as the major factor behind cancer anaemia and Vitamin D deficiency.

In our study, haematolymphoid malignancies had higher number of patients with underlying anaemia at presentation (71%).In absence of haematology department, majority of haematolymphoid malignancies are seen by medical oncology unit here, which in current study constitutes 19%(59/311) of all cases.^20,21^ Gall bladder is the most common cancer in this part of India.^22^ Gall bladder and gastrointestinal malignancies in our study, had 68% and 58% anaemia at presentation respectively. Compared to European studies, one of the reason for higher anaemia prevalence was presentation with more advanced stage, III and IV cancers (79%, 245/311) in our patient population, which reflects late presentation and aggressive biology^7,10,23.^

There were several limitations of our study. We only measured the laboratory values of haemoglobin, iron profile and serum B12 levels, but did not illicit the nature, type or severity of symptoms related to cancer anaemia such as fatigue, anorexia, breathlessness, palpitation or bleeding. As it was non interventional study, the decision to take iron or B12 supplementation was patient’s discretion and was not controlled, directed or followed by our team. Being a single point cross sectional study, we did not followed up patients longitudinally to study further drop in Haemoglobin with cancer directed therapies or extend of rise of haemoglobin and time taken to improve anaemic status after any supplementation. This is also the reason that we could not analyse treatment response, toxicities, outcomes or survival across several malignancies with respect to anaemia status at baseline.

Our study is a single largest institutional analysis reporting prevalence of anaemia in Eastern India, however this may not be a representative sample with respect to major metropolitan cities, West and South India. In our study target population was strictly defined as per the eligibility criteria, hence generalizability and external validity of our result beyond defined population cannot be guaranteed especially with respect to diverse geographical, temporal and ethical conditions. In a retrospective clinical audit done before finalising protocol for current study, it came to our notice that serum folate levels were not, reported consistently by our institutional laboratory. On further exploration we discovered that procurement and supply chain of folate assays were inconsistent due to several logistical issues leading to scarce reporting. Hence, we did not include serum folate measurements in our prospective study. Moreover, exclusive folate deficiency with normal B12 levels alone as cause of anaemia is seen in less than 2% of cancer patients.^16^ However, a separate comprehensive study is merited to evaluate its independent significance in treatment naive cancer patients with underlying anaemia.

## Conclusion

Anaemia is prevalent in sixty percent of treatment naive adult cancer patients. Iron deficiency, B12 deficiency and chronic inflammatory state are seen in two-third, half and one-third patients respectively. Identifying cause of anaemia at presentation, including nutritional deficiencies, may improve anaemic status after supplementation. Whether such pre-treatment probing of deficiencies and their subsequent correction impacts cancer outcomes and treatment toxicities favourably needs to be explored prospectively further.

## Data Availability

The authors confirm that the data supporting the findings of this study are available within the article

## Acknowledgements

1. Dr. Ruchi Kapoor, Oncquest Laboratories, New Delhi, India.

